# Brain-derived neurotrophic factor in cerebrospinal fluid and plasma is not a biomarker for Huntington’s disease

**DOI:** 10.1101/2020.11.02.20222166

**Authors:** Zhen-Yi Andy Ou, Lauren M. Byrne, Filipe B. Rodrigues, Rosanna Tortelli, Eileanoir B. Johnson, Martha S. Foiani, Marzena Arridge, Enrico De Vita, Rachael I. Scahill, Amanda Heslegrave, Henrik Zetterberg, Edward J. Wild

**Affiliations:** UCL Huntington’s Disease Centre, UCL Queen Square Institute of Neurology, University College London, London WC1N 3BG, UK; UK Dementia Research Institute at UCL, London WC1E 6BT, UK; Department of Neurodegenerative Disease, UCL Institute of Neurology, London WC1N 3BG, UK; Lysholm Department of Neuroradiology, National Hospital for Neurology and Neurosurgery, London WC1N 3BG, UK; Department of Biomedical Engineering, School of Biomedical Engineering and Imaging Sciences, King’s College London, London SE1 7EH, UK; Department of Psychiatry and Neurochemistry, Institute of Neuroscience and Physiology, the Sahlgrenska Academy at the University of Gothenburg, S-431 80 Mölndal, Sweden; Clinical Neurochemistry Laboratory, Sahlgrenska University Hospital, S-431 80 Mölndal, Sweden

## Abstract

Brain-derived neurotrophic factor (BDNF) is implicated in the survival of striatal neurons. BDNF function is reduced in Huntington’s disease (HD), possibly because mutant huntingtin impairs its cortico-striatal transport, contributing to striatal neurodegeneration. The BDNF trophic pathway is a therapeutic target, and blood BDNF has been suggested as a potential biomarker for HD, but BDNF has not been quantified in cerebrospinal fluid (CSF) in HD.

BDNF in CSF and plasma in the HD-CSF cohort (20 pre-manifest and 40 manifest HD mutation carriers and 20 age and gender-matched controls) were quantified using conventional ELISAs and an ultra-sensitive immunoassay.

BDNF concentration was below the limit of detection of the conventional ELISAs, raising doubt about previous CSF reports in neurodegeneration. Using the ultra-sensitive method, BDNF concentration was quantifiable in all samples but did not differ between controls and HD mutation carriers in CSF or plasma, was not associated with clinical scores or MRI brain volumetric measures, and had poor ability to discriminate controls from HD mutation carriers, and premanifest from manifest HD.

BDNF in CSF and plasma is unlikely to be a biomarker of HD progression, and urge caution in interpreting studies where conventional ELISA was used to quantify CSF BDNF.

## Introduction

Huntington’s disease (HD) is a fatal neurodegenerative disorder caused by a mutation in the gene encoding mutant huntingtin protein (mHTT). It causes behavioural, cognitive, and motor dysfunctions and no disease-modifying treatment has yet demonstrated efficacy^1^.

In HD, the selective vulnerability and degeneration of striatal neurons may be caused by the depletion of brain-derived neurotrophic factor (BDNF)^2^. BDNF is a growth factor implicated in neuronal survival, development, and synaptic plasticity. HD post-mortem brain tissue has been shown to have reduced BDNF^3^. Murine studies demonstrate that the absence of BDNF leads to early striatal neuronal death and HD phenotypes^4,5^. BDNF concentration is lower in the striatum of R6/1 and zQ175 mice compared to the wild-type counterparts^6,7^. This lowered concentration may have been influenced by a wild-type huntingtin-mediated mechanism. Wild-type huntingtin regulates the transport of BDNF and indirectly promotes the genetic expression of BDNF^8,9^ and BDNF overexpression produced phenotypic recovery in a murine model^10^. BDNF and its trophic pathway are targets for therapeutic development^11^.

BDNF concentration in cerebrospinal fluid (CSF) or blood could be an accessible means of quantifying dysfunction of this pathogenic pathway, could be a useful monitoring or prognosis biomarker, and could elucidate target engagement by therapeutics expected to restore the pathway. BDNF in these fluids has been studied in several neurodegenerative diseases: patients with Parkinson’s and Alzheimer’s disease (AD) have reduced serum BDNF levels^12,13^, and restoration of BDNF may ameliorate behavioural deficits and neuronal loss in AD models^12^. Similarly, BDNF deficiency in serum has been observed in HD patients^14^.

However, BDNF is stored in platelets^15^ so its concentration in blood may not be an accurate reflection of the CNS. Moreover, all previous reports of BDNF levels in CSF in neurodegeneration have used conventional enzyme-linked immunosorbent assays (ELISAs) and have found concentrations below the linear range of the assay, raising doubt as to the validity and accuracy of the reported disease differences^16–18^.

To our knowledge, BDNF has not been successfully quantified in CSF in HD patients^19^ and no comparison between CSF and blood levels has been performed using a suitable ultra-sensitive assay.

We therefore compared several immunoassays and quantified BDNF in blood and CSF from HD mutation carriers and healthy controls both cross-sectionally and over a 2-year longitudinal period to determine whether BDNF is a potential biomarker for HD.

## Materials and methods

### Study design

Participants (20 healthy controls, 20 premanifest HD (preHD), and 40 manifest HD) were recruited from the National Hospital for Neurology & Neurosurgery/University College London HD Multidisciplinary Clinic as part of a longitudinal CSF collection initiative (online protocol: https://doi.org/10.5522/04/11828448.v1)^20^. This was a single-site study affiliated with the HDClarity study(NCT02855476, http://hdclarity.net/) with added optional magnetic resonance imaging (MRI) and 6-week repeated sampling, and with a 24-month longitudinal follow-up. It was conducted according with the Declaration of Helsinki and was approved by the London–Camberwell St Giles Research Ethics Committee. All participants gave written informed consent.

### Study procedures

The inclusion and exclusion criteria, clinical assessments, blood and CSF collection and processing, and MRI acquisition and processing are described in the online protocol and previous reports^20,21^. Briefly, participants underwent clinical assessments including the Unified Huntington’s Disease Rating Scale (UHDRS) subscales. Blood samples were collected into lithium heparin tubes after an overnight fast and were centrifuged at 1300g for 10 minutes at 4°C to produce normal plasma (as opposed to platelet-poor plasma)^22^ and aliquoted on ice and stored at −80°C using a standardised protocol. T1-weighted MRI data were acquired on a 3T Siemens Prisma scanner as previously described. ^20^. Brain volume change was expressed as change per year and adjusted to total intracranial volume.

### Assay techniques

#### Immunoassays comparison

BDNF was quantified in duplicate in 20 CSF and serum samples from deidentified individuals from a clinical neurochemistry laboratory to compare the sensitivity of commercially available immunoassays: BDNF Emax ImmunoAssay System (Promega, USA), Human BDNF ELISA (Sigma-Aldrich, USA), and Single molecule array (Simoa) Human BDNF Discovery Kit (Quanterix, USA). POLARstar Omega (BMG Labtech, Germany) was used to measure the fluorescent products in the standard ELISAs, whilst an HD-1 Analyzer (Quanterix, USA) was used for the Simoa kit. For Promega and Sigma-Aldrich ELISAs, the limit of detection (LOD) and quantification (LOQ) were calculated from raw absorbance data.

#### The HD-CSF cohort

Plasma and CSF BDNF from the HD-CSF cohort were quantified in duplicate using the Simoa kit on an HD-1 Analyzer according to manufacturer guidance. Plasma samples were measured at 1:640 and CSF at 1:4 dilution. The lower LOQ and lower LOD were 0.0293 pg/mL and 0.0026 pg/mL, respectively. All measurements were above both. The intra-assay coefficients of variation (CV; calculated as the mean of the CVs for each sample’s duplicate measurements) was 8.30% for CSF (n=84) and 3.26% for plasma (n=91). The inter-assay CV (calculated as the mean of the CVs for analogous spiked positive controls provided by the manufacturer and used in each well plate) was 4.39% for CSF (n=2) and 2.74% for plasma (n=4). Four (4.8%) CSF samples were only measured once. Quantification blinded to disease status.

### Statistical methods

For the HD-CSF cohort, BDNF distribution was assessed for normality, and plasma and CSF BDNF values underwent square root and inverse square transformation, respectively.

General linear models and Pearson’s chi-squared or Fisher’s exact test were performed to assess intergroup differences at baseline and at the 24-month follow-up characteristics.

Potential confounders including age, gender, body mass index (BMI), medication, serum platelet counts, and CSF blood contamination (hemoglobin and erythrocytes), storage duration, and short problem behaviour assessment (PBA-s) subscores were examined through general linear models, Pearson’s correlations or ordinal logistic regressions. Those expected to have a relevant impact on BDNF, either based on reports from the literature, or directly seen in our sample, were included as covariates for subsequent models^15,23–26^. All included gender, BMI, anti-depressant, anti-psychotic medication, age. For plasma we also included sample storage duration, and platelet count. For CSF we also included CSF erythrocyte count. As a relevant contributor to HD natural history, CAG repeat length was included in the models.

BDNF intergroup differences were investigated with general linear models. Association in HD mutation carriers with clinical and imaging measures were investigated with partial Pearson’s correlations with bootstrapped bias corrected and accelerated 95% confidence intervals (95%CI). Receiver operating characteristic (ROC) curves were drawn for plasma and CSF BDNF.

To investigate short term stability, two-way mixed effects models intraclass correlations (ICC) were performed between the baseline and optional 6-week samples.

Annualised rates of change in BDNF and clinical measures were calculated by subtracting baseline from follow-up value and dividing by time between samplings. Intergroup differences and associations in HD mutation carriers with clinical and imaging measures were investigated as above.

To study BDNF longitudinal trajectories, we used mixed effects models with age and potential confounders as fixed effects, and random effects for participant (intercept) and age (slope), generated independently for controls and mutation carriers.

All analyses were performed with Stata 15.1 (StataCorp). The significance level was defined as p-value < 0.05.

### Data availability

The data that support the findings of this study are available from the corresponding author, EJW, upon reasonable request.

## Results

### 1. Immunoassay comparison

Serum BDNF levels in 20 test samples were in the linear range for tested assays. However, CSF levels were below the LOD in all 20 samples on the Promega assay, while 16 (80%) out of 20 samples were between the LOD and the LOQ on the Sigma-Aldrich assay. In plasma, all tested assays were correlated (Simoa and Promega: r=0.9200, P<0.0001; Simoa and Sigma-Aldrich: r=0.9428, P<0.0001; Promega and Sigma-Aldrich: r=0.9175, P<0.0001). In CSF, Simoa and Sigma-Aldrich showed a weaker association (r=0.4269, P=0.0991). The Simoa assay quantified BDNF concentration in blood and CSF within the linear range, and above the LOD and LOQ in all samples. This assay was therefore used for all subsequent analyses in both biofluids.

### 2. Cross-sectional analysis of baseline

#### 2.1 Demographic characteristics and confounding variables

Demographic and clinical characteristics are summarised in Table 1. The control group was matched to the HD mutation carriers as a whole, but was older on average than preHD, which in turn was younger than the HD group. The preHD group had a lower mean BMI than the controls, but did not differ from HD. More manifest HD participants were on medication, including anti-depressants and anti-psychotics.

**Table 1.**
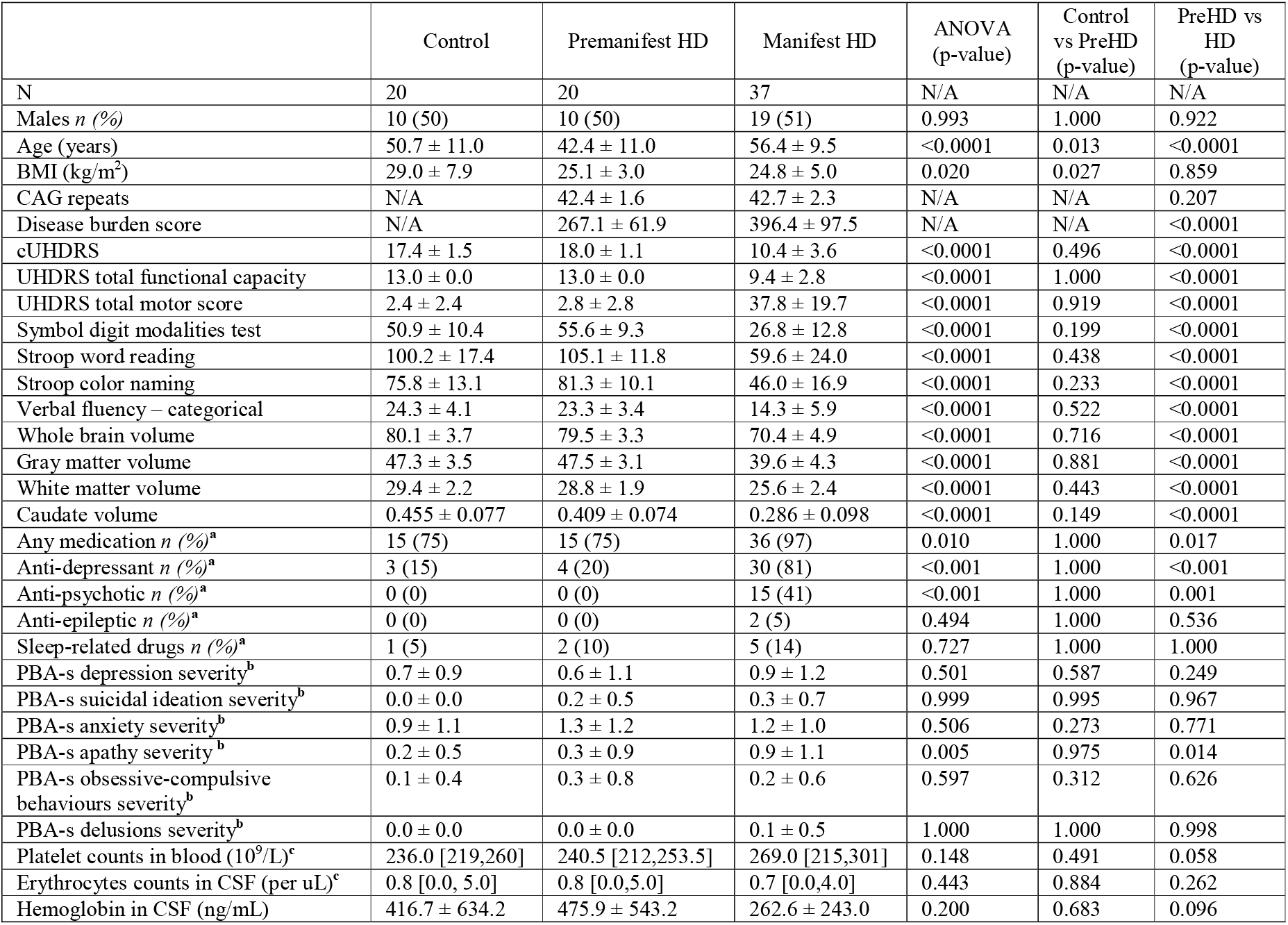
Basic characteristics of the baseline HD-CSF. Presented p values are not adjusted for multiple comparisons. Results were replicated at follow-up. Brain volumes are percentages of total intracranial volume. Values are mean ± SD, except where it stated otherwise. Manifest HD, manifest HD mutation carriers; Premanifest HD/preHD, premanifest HD mutation carriers; SD, standard deviation; ANOVA, analysis of variance; cUHDRS, composite Unified Huntington’s Disease Ratings Scale; PBA-s, Problem Behaviours Assessment-short version; CSF, cerebrospinal fluid; BMI, body-mass index; cUHDRS, composite Unified Huntington’s Disease Ratings Scale; UHDRS, Unified Huntington’s Disease Ratings Scale; N/A, not applicable. ^**a**^ Fisher’s exact test; ^**b**^ ordinal logistic regression; ^**c**^ median [Q1, Q3].

|  | Control | Premanifest HD | Manifest HD | ANOVA<br>(p-value) | Control<br>vs PreHD<br>(p-value) | PreHD vs<br>HD<br>(p-value) |
| --- | --- | --- | --- | --- | --- | --- |
| N | 20 | 20 | 37 | N/A | N/A | N/A |
| Males *n* (%) | 10 (50) | 10 (50) | 19 (51) | 0.993 | 1.000 | 0.922 |
| Age (years) | 50.7 ± 11.0 | 42.4 ± 11.0 | 56.4 ± 9.5 | <0.0001 | 0.013 | <0.0001 |
| BMI (kg/m<sup>2</sup>) | 29.0 ± 7.9 | 25.1 ± 3.0 | 24.8 ± 5.0 | 0.020 | 0.027 | 0.859 |
| CAG repeats | N/A | 42.4 ± 1.6 | 42.7 ± 2.3 | N/A | N/A | 0.207 |
| Disease burden score | N/A | 267.1 ± 61.9 | 396.4 ± 97.5 | N/A | N/A | <0.0001 |
| cUHDRS | 17.4 ± 1.5 | 18.0 ± 1.1 | 10.4 ± 3.6 | <0.0001 | 0.496 | <0.0001 |
| UHDRS total functional capacity | 13.0 ± 0.0 | 13.0 ± 0.0 | 9.4 ± 2.8 | <0.0001 | 1.000 | <0.0001 |
| UHDRS total motor score | 2.4 ± 2.4 | 2.8 ± 2.8 | 37.8 ± 19.7 | <0.0001 | 0.919 | <0.0001 |
| Symbol digit modalities test | 50.9 ± 10.4 | 55.6 ± 9.3 | 26.8 ± 12.8 | <0.0001 | 0.199 | <0.0001 |
| Stroop word reading | 100.2 ± 17.4 | 105.1 ± 11.8 | 59.6 ± 24.0 | <0.0001 | 0.438 | <0.0001 |
| Stroop color naming | 75.8 ± 13.1 | 81.3 ± 10.1 | 46.0 ± 16.9 | <0.0001 | 0.233 | <0.0001 |
| Verbal fluency – categorical | 24.3 ± 4.1 | 23.3 ± 3.4 | 14.3 ± 5.9 | <0.0001 | 0.522 | <0.0001 |
| Whole brain volume | 80.1 ± 3.7 | 79.5 ± 3.3 | 70.4 ± 4.9 | <0.0001 | 0.716 | <0.0001 |
| Gray matter volume | 47.3 ± 3.5 | 47.5 ± 3.1 | 39.6 ± 4.3 | <0.0001 | 0.881 | <0.0001 |
| White matter volume | 29.4 ± 2.2 | 28.8 ± 1.9 | 25.6 ± 2.4 | <0.0001 | 0.443 | <0.0001 |
| Caudate volume | 0.455 ± 0.077 | 0.409 ± 0.074 | 0.286 ± 0.098 | <0.0001 | 0.149 | <0.0001 |
| Any medication *n* (%)<sup>a</sup> | 15 (75) | 15 (75) | 36 (97) | 0.010 | 1.000 | 0.017 |
| Anti-depressant *n* (%)<sup>a</sup> | 3 (15) | 4 (20) | 30 (81) | <0.001 | 1.000 | <0.001 |
| Anti-psychotic *n* (%)<sup>a</sup> | 0 (0) | 0 (0) | 15 (41) | <0.001 | 1.000 | 0.001 |
| Anti-epileptic *n* (%)<sup>a</sup> | 0 (0) | 0 (0) | 2 (5) | 0.494 | 1.000 | 0.536 |
| Sleep-related drugs *n* (%)<sup>a</sup> | 1 (5) | 2 (10) | 5 (14) | 0.727 | 1.000 | 1.000 |
| PBA-s depression severity<sup>b</sup> | 0.7 ± 0.9 | 0.6 ± 1.1 | 0.9 ± 1.2 | 0.501 | 0.587 | 0.249 |
| PBA-s suicidal ideation severity<sup>b</sup> | 0.0 ± 0.0 | 0.2 ± 0.5 | 0.3 ± 0.7 | 0.999 | 0.995 | 0.967 |
| PBA-s anxiety severity<sup>b</sup> | 0.9 ± 1.1 | 1.3 ± 1.2 | 1.2 ± 1.0 | 0.506 | 0.273 | 0.771 |
| PBA-s apathy severity<sup>b</sup> | 0.2 ± 0.5 | 0.3 ± 0.9 | 0.9 ± 1.1 | 0.005 | 0.975 | 0.014 |
| PBA-s obsessive-compulsive behaviours severity<sup>b</sup> | 0.1 ± 0.4 | 0.3 ± 0.8 | 0.2 ± 0.6 | 0.597 | 0.312 | 0.626 |
| PBA-s delusions severity<sup>b</sup> | 0.0 ± 0.0 | 0.0 ± 0.0 | 0.1 ± 0.5 | 1.000 | 1.000 | 0.998 |
| Platelet counts in blood (10<sup>9</sup>/L)<sup>c</sup> | 236.0 [219,260] | 240.5 [212,253.5] | 269.0 [215,301] | 0.148 | 0.491 | 0.058 |
| Erythrocytes counts in CSF (per uL)<sup>c</sup> | 0.8 [0.0, 5.0] | 0.8 [0.0,5.0] | 0.7 [0.0,4.0] | 0.443 | 0.884 | 0.262 |
| Hemoglobin in CSF (ng/mL) | 416.7 ± 634.2 | 475.9 ± 543.2 | 262.6 ± 243.0 | 0.200 | 0.683 | 0.096 |

Plasma and CSF BDNF did not differ by gender (Figure S1). The platelet count in blood did not differ between groups, but it was positively associated with plasma BDNF in HD mutation carriers (Figure S2). In the HD group, the average erythrocyte count was negatively correlated with CSF BDNF concentration (Figure S2).

#### 2.2 Storage duration as a BDNF confounder in plasma but not CSF

BDNF levels in baseline plasma samples were substantially higher than in the 24-month follow-up samples (Figure 1). This was not seen in CSF, suggesting the possibility that BDNF levels were rising artefactually over time in frozen plasma, perhaps due to leakage from small numbers of residual platelets or degradation of pro-BDNF to BDNF. In keeping with this, the time spent in frozen storage was positively associated with plasma BDNF but not CSF BDNF, in controls and HD mutation carriers (Figure 1). Hence, plasma BDNF analyses were additionally adjusted for sample storage duration.

**Figure 1.**
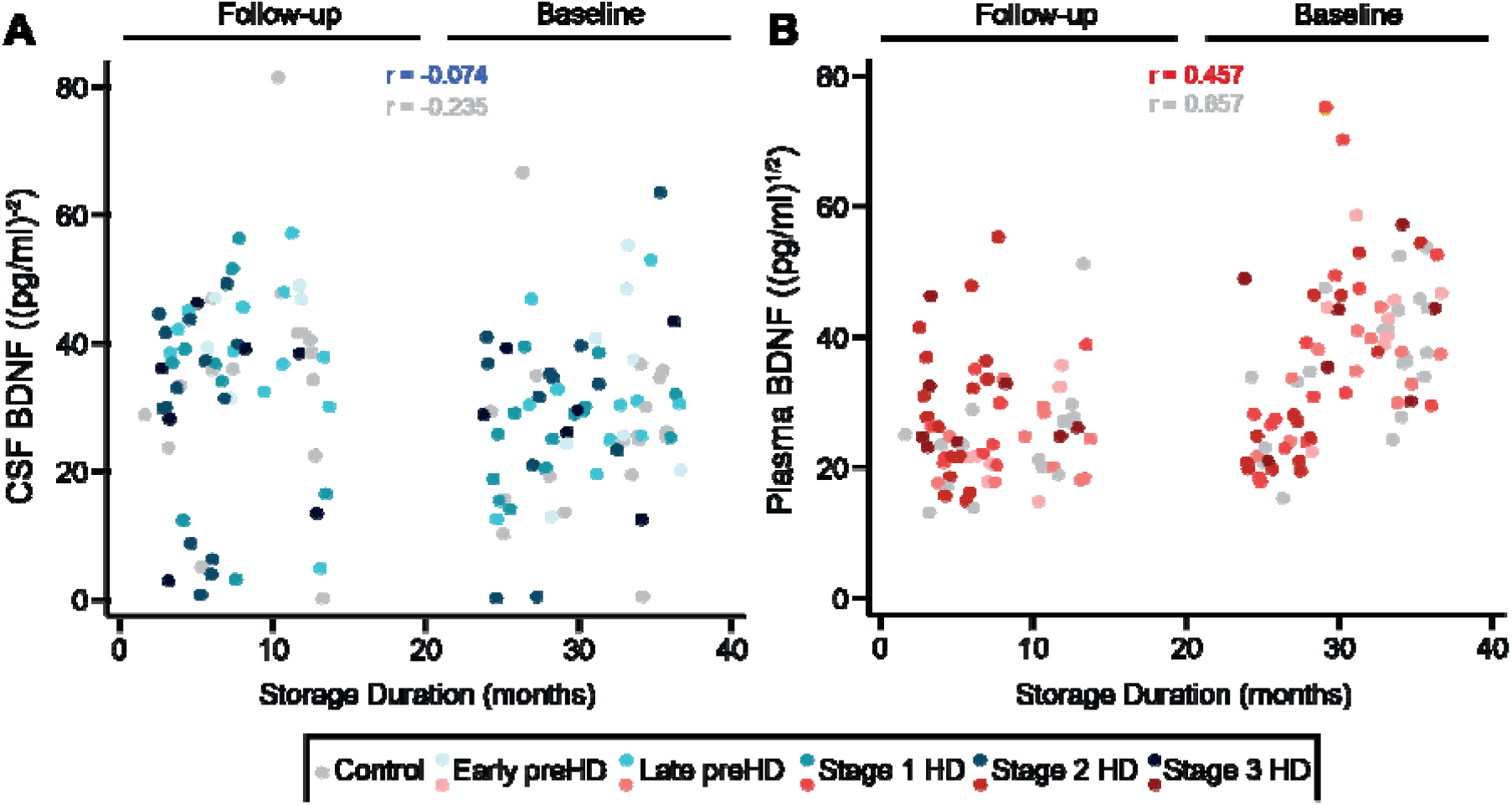
Association between biofluid storage duration and BDNF concentrations. Correlation between the storage duration in months and CSF BDNF (A) and plasma BDNF (B). CSF BDNF values were inverse square transformed and plasma BDNF were square root transformed. CSF, cerebrospinal fluid; HD, manifest HD mutation carriers; PreHD, premanifest HD mutation carriers; BDNF, brain-derived neurotrophic factor.

#### 2.3 BDNF levels in Huntington’s disease

Neither plasma nor CSF BDNF differed significantly between groups (Figure 2). There were no associations between plasma or CSF BDNF and clinical measures, with the sole exception of plasma BDNF which was weakly positively associated with Verbal Fluency - Categorical (r=0.261, p=0.015; Figure S3). There were no associations between BDNF and imaging measures, except a weak positive association between CSF BDNF and white matter volume (r=0.306, p=0.032; Figure S4). It is of note that no correction for multiplicity was applied.

**Figure 2.**
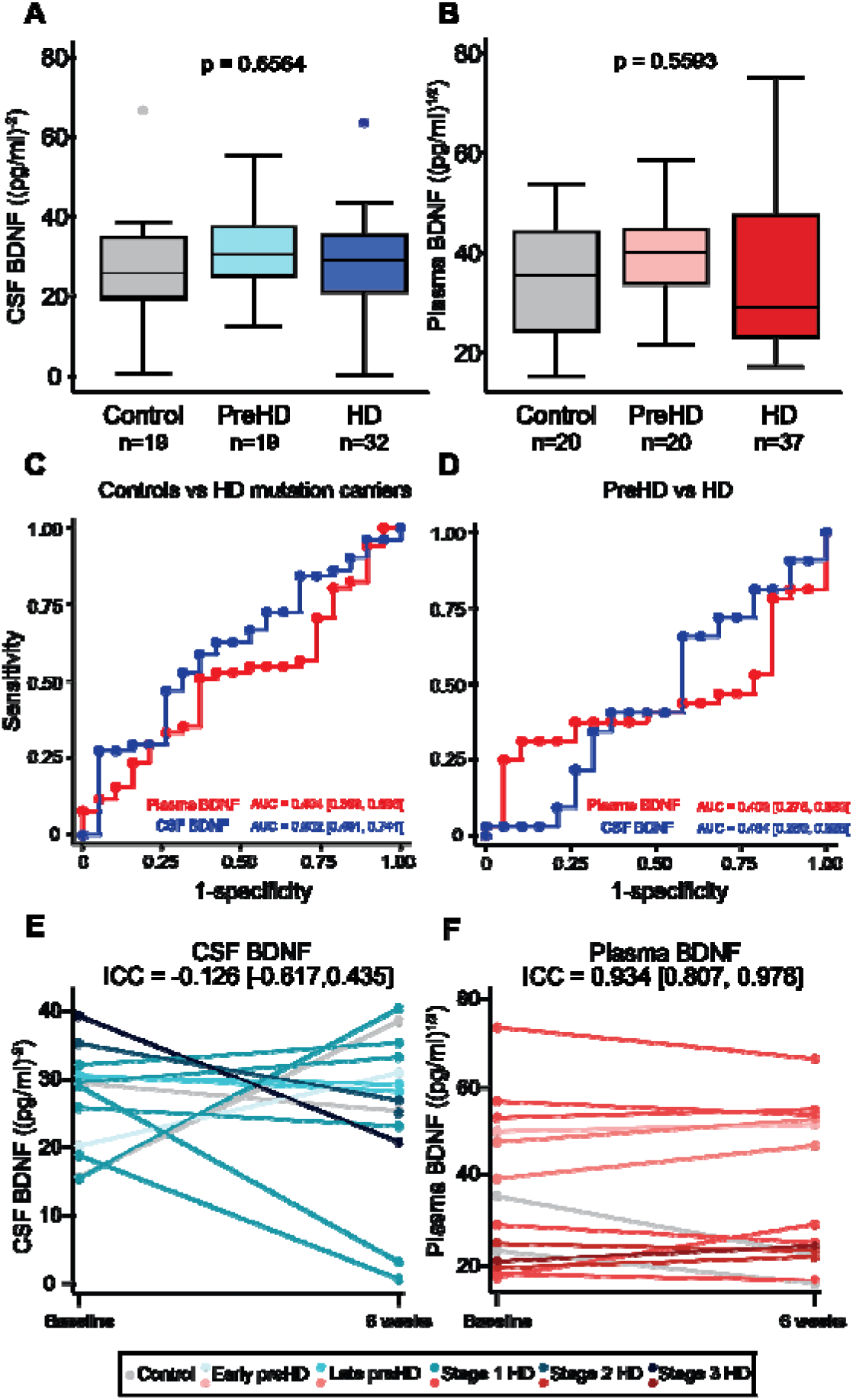
Baseline BDNF plots. Concentration of BDNF in CSF (A) and plasma (B) in healthy controls, premanifest HD (PreHD), and manifest HD (HD) patients. Comparison were generated with general linear models and were adjusted for CSF (gender, BMI, anti-depressant, anti-psychotic medication, age, CAG repeats, and erythrocyte count) or plasma covariates (gender, BMI, anti-depressant, anti-psychotic medication, age, CAG repeats, sample storage duration, and platelet count). ROC curves for (C) discrimination between healthy control (n=19) and HD mutation carriers (n=51) and (D) discrimination between premanifest HD (n=19) and HD patients (n=32). Results A-D were replicated at follow-up. Stability of BDNF in CSF (E) and plasma (F) over approximately 6 weeks was assessed with two-way mixed effects models intraclass correlation (ICC). Each line described the same participant (n=13 in CSF and n=14 in plasma) between the baseline and optional 6-week repeated sampling. CSF BDNF values were inverse square transformed and plasma BDNF were square root transformed. CSF, cerebrospinal fluid; preHD, premanifest HD mutation carriers; HD, manifest HD mutation carriers; BDNF, brain-derived neurotrophic factor; ROC, receiver operating characteristic; BMI, body-mass index; CAG, cytosine-adenine-guanine; ICC, intraclass correlation.

Plasma and CSF BDNF showed poor ability to distinguish controls from HD mutation carriers (plasma AUC=0.494, CSF AUC=0.602; Figure 2C) and preHD from HD (plasma AUC 0.409, CSF AUC=0.454; Figure 2D).

CSF BDNF concentrations significantly fluctuated between the baseline and 6-week follow-up period (CSF ICC=-0.126, Figure 2E). Plasma concentrations were stable (ICC=0.934, Figure 2F).

### 3. Longitudinal analysis

The rate of change in CSF BDNF did not differ between any HD stages (Figure 3A-B). Longitudinal evaluation of plasma BDNF was limited by its apparent tendency to rise in freezer storage, producing spuriously higher values for baseline samples (Figure 3C-D), but using the mixed effect model with adjustment for confounders, CSF and plasma BDNF did not change over time in controls or HD mutation carriers (Figure 3E-F). Overall, baseline BDNF did not predict subsequent change in clinical measures (Figure S5), nor did the rates of change in BDNF (Figure S6).

**Figure 3.**
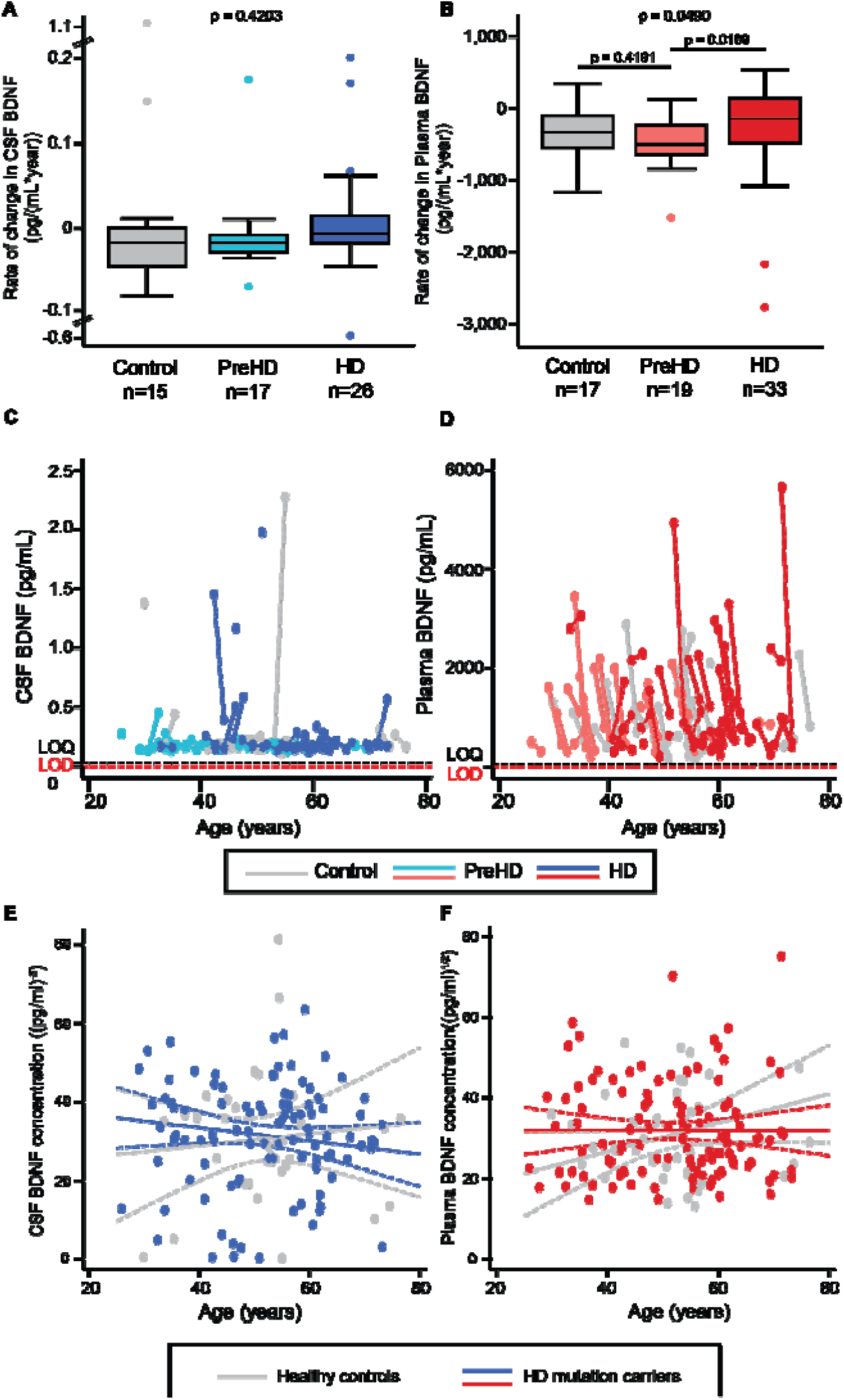
Longitudinal analyses in CSF and plasma. Rate of change in the concentration of BDNF in CSF (A) and plasma (B) in healthy controls, premanifest HD (PreHD), and manifest HD (HD) patients. Comparison were generated with general linear models. Relationship between raw CSF (C) and plasma BDNF values (D) and age in years. The limit of quantification (LOQ) and limit of detection (LOD) of the SIMOA assay are indicated. Longitudinal trajectories were studied with mixed effect models, which are adjusted for covariates (E,F). CSF, cerebrospinal fluid; BDNF, brain-derived neurotrophic factor; HD, manifest HD mutation carriers, PreHD, premanifest HD mutation carriers.

## Discussion

The conventional ELISAs we tested lacked the sensitivity required to accurately quantify CSF BDNF. With an ultra-sensitive assay, BDNF levels were measurable in both CSF and plasma but did not reflect HD or its progression and were not associated with the examined clinical and imaging measures.

It was notable that the ELISA assays used in previous reports of altered CSF BDNF in neurological disease appear insufficiently sensitive for the purpose. This emphasizes the importance of ensuring the analyte in question lies in the assay’s linear range and urges caution when interpreting previous reports of CSF BDNF as a disease biomarker. To our knowledge, this is therefore the first report of BDNF levels in human CSF using an assay capable of reliably quantifying them.

Unexpectedly, we also found that the storage duration is the major influence upon BDNF level in plasma but not CSF. This diminishes the utility of stored frozen samples for BDNF quantification in blood products and suggests time in storage should be controlled for in future biomarker studies. BDNF is known to be enriched in platelets, and our centrifugation procedure was designed to produce normal as opposed to platelet-poor plasma. The latter may show less influence from platelet-derived BDNF and therefore be a better surrogate for brain BDNF^22^

BDNF had no properties suggesting it is a biomarker for HD progression in CSF or plasma. Its level did not differ significantly between controls and HD mutation carriers, or between preHD and HD groups. It had poor classification ability in our ROC analysis and lacked associations with clinical and MRI brain volumetric measures. In the context of a clear lack of groupwise differences, the small number of associations seen among many tests is likely a chance finding and not of biological significance. Neither baseline BDNF level, nor its rate of change, were prognostic for subsequent change in clinical or imaging outcomes. We therefore conclude that BDNF in CSF or plasma is unlikely to be a valid biomarker of HD, in the ‘diagnostic’, ‘monitoring’ or ‘prognostic’ categories according to the FDA’s BEST categorisation.

These negative findings contrast starkly with the performance of other CSF and plasma analytes, most notably neurofilament light and mutant huntingtin proteins, which do differ between groups, rise with progression, and predict subsequent clinical and imaging change^20,21,27^.

It should be noted that the absence of detectable changes in CSF or plasma does not exclude an important role for BDNF or its trophic pathway in the pathogenesis of HD. The decrease of BDNF might be very region-specific and therefore overwhelmed by generally unaltered BDNF in other regions. Several studies found reduced BDNF in striatum but not cortex in HD models^3,7,9^. Moreover, if striatal BDNF loss is significant, other promotors of the BDNF gene that are not directly modulated by HTT could possibly compensate for the mHTT-induced deficit of BDNF in the cortex^28^.

In the present study, we did not investigate BDNF isoforms, including its precursor, pro-BDNF. Studying the isoforms of BDNF, including pro-BDNF, could still reveal disease-related changes. Unlike some previous reports, we studied plasma rather than serum, but since plasma is prepared by centrifugation rather than clotting, we reasoned it was less likely to contain artefactual signals from platelets.

Finally, it is important to note that even though we found BDNF to lack utility as a biomarker of HD in the natural history setting, it is still theoretically possible that a therapeutic intervention could produce detectable and meaningful changes in CSF or plasma BDNF. Our findings may be of value for the design of such trials in which BDNF would serve as a pharmacodynamic biomarker.

## Supporting information

STROBE

## Acknowledgements

We would like to thank all the participants from the HD community who donated samples and gave their time to take part in this study.

## Funding

This work was supported by the Medical Research Council UK, the CHDI foundation, the Huntington’s Disease Society of America, the Hereditary Disease Foundation, the Wellcome Trust (Wellcome Collaborative Award In Science 200181/Z/15/Z and Wellcome/EPSRC Centre for Medical Engineering [WT 203148/Z/16/Z]), the Department of Health’s NIHR Biomedical Research Centres funding scheme, F. Hoffmann-La Roche Ltd, Horizon 2020 Framework Programme, the Swedish Research Council, the European Research Council, and Swedish State Support for Clinical Research, and the Innovative Medicines Initiative Joint Undertaking under EMIF grant. HZ is a Wallenberg Scholar.

## Author contributions

EJW designed the study with the input of HZ and RIS. FBR and LMB were involved in participant recruitment. Eligibility, clinical examinations and sample collection were performed by FBR, LBM, and RT. Imaging assessments were conceived RIS, EBJ and EDV, data was acquired by EDV, MA, EBJ, and processed by RIS and EBJ. ZAO, MF and LMB processed and analysed the patient samples. FBR developed and ZAO performed the statistical analysis. ZAO, FBR, LMB and EJW interpreted the data and wrote the manuscript; and all authors contributed to reviewing the manuscript.

## Competing interests

ZAO, LMB, FBR, MF, RT, EBJ, RIS, AH, HZ, EJW are University College London employees. MA is a University College London Hospitals NHS Foundation Trust employee. EDV is a King’s College London employee. LMR has provided consultancy services to GLG, F. Hoffmann-La Roche Ltd, Genentech and Annexon. FBR has provided consultancy services to GLG and F. Hoffmann-La Roche Ltd. RIS has undertaken consultancy services for Ixitech Ltd. HZ has served at scientific advisory boards for Denali, Roche Diagnostics, Wave, Samumed, Siemens Healthineers, Pinteon Therapeutics and CogRx, has given lectures in symposia sponsored by Fujirebio, Alzecure and Biogen, and is a co-founder of Brain Biomarker Solutions in Gothenburg AB (BBS), which is a part of the GU Ventures Incubator Program (outside submitted work). EJW reports grants from Medical Research Council, CHDI Foundation, and F. Hoffmann-La Roche Ltd during the conduct of the study; personal fees from Hoffman La Roche Ltd, Triplet Therapeutics, PTC Therapeutics, Shire Therapeutics, Wave Life Sciences, Mitoconix, Takeda and Loqus23. All honoraria for these consultancies were paid through the offices of UCL Consultants Ltd., a wholly owned subsidiary of University College London. University College London Hospitals NHS Foundation Trust has received funds as compensation for conducting clinical trials for Ionis Pharmaceuticals, Pfizer and Teva Pharmaceuticals.

## Notes

### Clinical Protocols

https://doi.org/10.5522/04/11828448.v1

### Author Declarations

It was conducted according with the Declaration of Helsinki and was approved by the London-Camberwell St Giles Research Ethics Committee. All participants gave written informed consent.

## References

1. Rodrigues FB, Abreu D, Damásio J, et al. Survival, Mortality, Causes and Places of Death in a European Huntington’s Disease Prospective Cohort. Mov Disord Clin Pract. 2017;4:737–742.

2. Zuccato C, Ciammola A, Rigamonti D, et al. Loss of huntingtin-mediated BDNF gene transcription in Huntington’s disease. Science (80-) [online serial]. 2001;293:493–498.

3. Ferrer I, Goutan E, Marin C, Rey M., Ribalta T. Brain-derived neurotrophic factor in Huntington disease. Brain Res [online serial]. I. Ferrer, Unitat de Neuropatologia, Servei d’Anatomia Patologica, Hospital princeps d’Espanya, Barcelona, Spain.: Elsevier (P.O. Box 211, Amsterdam 1000 AE, Netherlands); 2000;866:257–261.

4. Rauskolb S, Zagrebelsky M, Dreznjak A, et al. Global Deprivation of Brain-Derived Neurotrophic Factor in the CNS Reveals an Area-Specific Requirement for Dendritic Growth. J Neurosci. Epub 2010.

5. Strand AD, Baquet ZC, Aragaki AK, et al. Expression profiling of Huntington’s disease models suggests that brain-derived neurotrophic factor depletion plays a major role in striatal degeneration. J Neurosci [online serial]. A.D. Strand, Fred Hutchinson Cancer Research Center, 1100 Fairview Avenue North, Seattle, WA 98109, United States.: Society for Neuroscience (1121 14th Street,NW, Suite 1010, Washington DC 20005, United States); 2007;27:11758–11768.

6. Ma Q, Yang J, Li T, Milner TA, Hempstead BL. Selective reduction of striatal mature BDNF without induction of proBDNF in the zQ175 mouse model of Huntington’s disease. Neurobiol Dis. Epub 2015.

7. Gharami K, Xie Y, An JJ, Tonegawa S, Xu B. Brain-derived neurotrophic factor over- expression in the forebrain ameliorates Huntington’s disease phenotypes in mice. J Neurochem. Epub 2008.

8. Zuccato C, Belyaev N, Conforti P, et al. Widespread Disruption of Repressor Element- 1 Silencing Transcription Factor/Neuron-Restrictive Silencer Factor Occupancy at Its Target Genes in Huntington’s Disease. J Neurosci. Epub 2007.

9. Gauthier LR, Charrin BC, Borrell-Pagès M, et al. Huntingtin Controls Neurotrophic Support and Survival of Neurons by Enhancing BDNF Vesicular Transport along Microtubules. Cell [online serial]. S. Humbert, Unite Mixte de Recherche 146, Ctr. Natl. de la Rech. Scientifique, Centre Universitaire, 91405 Orsay Cedex, France. E- mail:: Cell Press (1100 Massachusetts Avenue, Cambridge MA 02138-9957, United States); 2004;118:127–138.

10. Xie Y, Hayden MR, Xu B. BDNF Overexpression in the Forebrain Rescues Huntington’s Disease Phenotypes in YAC128 Mice. J Neurosci. Epub 2010.

11. Todd D, Gowers I, Dowler SJ, et al. A monoclonal antibody TrkB receptor agonist as a potential therapeutic for huntington’s disease. PLoS One [online serial]. CHDI Management, CHDI Foundation, Princeton, NJ, United States: Public Library of Science (185 Berry Street, Suite 1300, San Francisco CA 94107, United States); 2014;9:e87923.

12. Jiao SS, Shen LL, Zhu C, et al. Brain-derived neurotrophic factor protects against tau-related neurodegeneration of Alzheimer’s disease. Transl Psychiatry. Epub 2016.

13. Scalzo P, Kümmer A, Bretas TL, Cardoso F, Teixeira AL. Serum levels of brain-derived neurotrophic factor correlate with motor impairment in Parkinson’s disease. J Neurol. Epub 2010.

14. Ciammola A, Sassone J, Cannella M, et al. Low brain-derived neurotrophic factor (BDNF) levels in serum of Huntington’s disease patients. Am J Med Genet Part B Neuropsychiatr Genet [online serial]. A. Ciammola, Department of Neurology, Laboratory of Neuroscience, IRCCS Istituto Auxologico Italiano, via Spagnoletto 3, 20149 - Milan, Italy.: Wiley-Liss Inc. (111 River Street, Hoboken NJ 07030-5774, United States); 2007;144:574–577.

15. Fujimura H, Altar CA, Chen R, et al. Brain-derived neurotrophic factor is stored in human platelets and released by agonist stimulation. Thromb Haemost. Epub 2002.

16. Burbach GJ, Hellweg R, Haas CA, Del Turco D, Deicke U, Abramowski D. Induction of Brain-Derived Neurotrophic Factor in Plaque-Associated Glial Cells of Aged APP23 Transgenic Mice. J Neurosci [online serial]. 2004;24:2421–2430.

17. Kizawa-Ueda M, Ueda A, Kawamura N, et al. Neurotrophin levels in cerebrospinal fluid of adult patients with meningitis and encephalitis. Eur Neurol [online serial]. 2011;65:138–143.

18. Laske C, Stransky E, Leyhe T, et al. BDNF serum and CSF concentrations in Alzheimer’s disease, normal pressure hydrocephalus and healthy controls. J Psychiatr Res [online serial]. 2007;41:387–394.

19. Rodrigues FB, Byrne LM, Wild EJ. Biofluid biomarkers in Huntington’s disease. Methods Mol Biol. 2018. p. 329–396.

20. Byrne LM, Rodrigues FB, Johnson EB, et al. Evaluation of mutant huntingtin and neurofilament proteins as potential markers in Huntington’s disease. Sci Transl Med. Epub 2018.

21. Rodrigues FB, Byrne LM, Tortelli R, et al. Longitudinal dynamics of mutant huntingtin and neurofilament light in Huntington’s disease: the prospective HD-CSF study. medRxiv [online serial]. Epub 2020.:2020.03.31.20045260.

22. Gejl AK, Enevold C, Bugge A, Andersen MS, Nielsen CH, Andersen LB. Associations between serum and plasma brain-derived neurotrophic factor and influence of storage time and centrifugation strategy. Sci Rep. 2019;9.

23. Pillai A, Bruno D, Sarreal AS, et al. Plasma BDNF levels vary in relation to body weight in females. PLoS One. Epub 2012.

24. Jung SH, Kim J, Davis JM, Blair SN, Cho HC. Association among basal serum BDNF, cardiorespiratory fitness and cardiovascular disease risk factors in untrained healthy Korean men. Eur J Appl Physiol. Epub 2011.

25. Li G, Peskind ER, Millard SP, et al. Cerebrospinal fluid concentration of brain-derived neurotrophic factor and cognitive function in non-demented subjects. PLoS One. Epub 2009.

26. Lommatzsch M, Zingler D, Schuhbaeck K, et al. The impact of age, weight and gender on BDNF levels in human platelets and plasma. Neurobiol Aging. Epub 2005.

27. Byrne LM, Rodrigues FB, Blennow K, et al. Neurofilament light protein in blood as a potential biomarker of neurodegeneration in Huntington’s disease: a retrospective cohort analysis. Lancet Neurol. Epub 2017.

28. Maynard KR, Hill JL, Calcaterra NE, et al. Functional Role of BDNF Production from Unique Promoters in Aggression and Serotonin Signaling. Neuropsychopharmacology. 2016;41:1943–1955.

