## Supplementary Materials for "Brain-derived neurotrophic factor in cerebrospinal fluid and plasma is not a biomarker for Huntington’s disease"

### Sample size

An *a priori* power calculation was performed for the HD-CSF study based on CSF analysis of mHTT through the usage of single molecule counting immunoassay, which was outlined in the HD-CSF protocol^1^. Twenty subjects per group gives >90% power at 5% significance level to detect predicted changes.

### Study procedures

#### CSF and blood sampling and processing

CSF and blood collection were performed between 8:00 and 10:30 AM local time (London, United Kingdom). CSF was gathered through lumbar puncture protocol and plasma collection was performed in lithium heparin tubes. All participants were requested to fast from midnight before their appointment. Please refer to the published protocol, as previously described ^2^. Platelet count in blood was quantified at the screening visit, up to 30 day before CSF and plasma sampling visits.

#### MRI acquisition and processing

3D volume T1-weighted MRI data were acquired on a 3T Siemens Prisma scanner. MIDAS, SPM12 “Segment”, and MALP-EM were used to process the MRI data in order to measure the volume of whole-brain, gray matter, white matter, and caudate. Please refer to HD-CSF study for MRI acquisition and processing protocol, available at <https://doi.org/10.5522/04/11828448.v1>^2^. Follow-up caudate volume was computed by the baseline volume minus the amount of atrophy measured via the caudate boundary shift integral, and follow-up grey and white matter volumes were calculated by subtracting the amount of atrophy from baseline volumes.

#### Clinical assessments

Motor, cognitive and functional status were assessed using the UHDRS from the core Enroll-HD battery, including: the UHDRS Total Motor Score, Total Functional Capacity, Symbol Digit Modalities Test, Stroop Word Reading, Stroop Color Naming, Verbal Fluency – Categorical and Problem Behaviours Assessment short form (PBA-s). These were performed at either the screening or an associated Enroll-HD visit (https://www.enroll-hd.org) within the 2 months prior to screening. We employed a calibrated iteration of the composite UHDRS (cUHDRS)^3^. The cUHDRS was chosen as the primary outcome measure for the analysis of clinical progression as it has favourable signal-to-noise characteristics, encompasses clinical deterioration across multiple relevant domains and has regulatory acceptance as a meaningful measure of HD severity.

### References:

1.	Wild EJ, Boggio R, Langbehn D, et al. Quantification of mutant huntingtin protein in cerebrospinal fluid from Huntington’s disease patients. J Clin Invest. Epub 2015.

2.	Byrne LM, Rodrigues FB, Johnson EB, et al. Evaluation of mutant huntingtin and neurofilament proteins as potential markers in Huntington’s disease. Sci Transl Med. Epub 2018.

3.	Trundell D, Palermo G, Schobel S, Long JD, Leavitt BR, Tabrizi SJ. F23 Validity, reliability, ability to detect change and meaningful within-patient change of the CUHDRS. J Neurol Neurosurg Psychiatry [online serial]. 2018;89:A48 LP-A48.

### Supplementary Figures

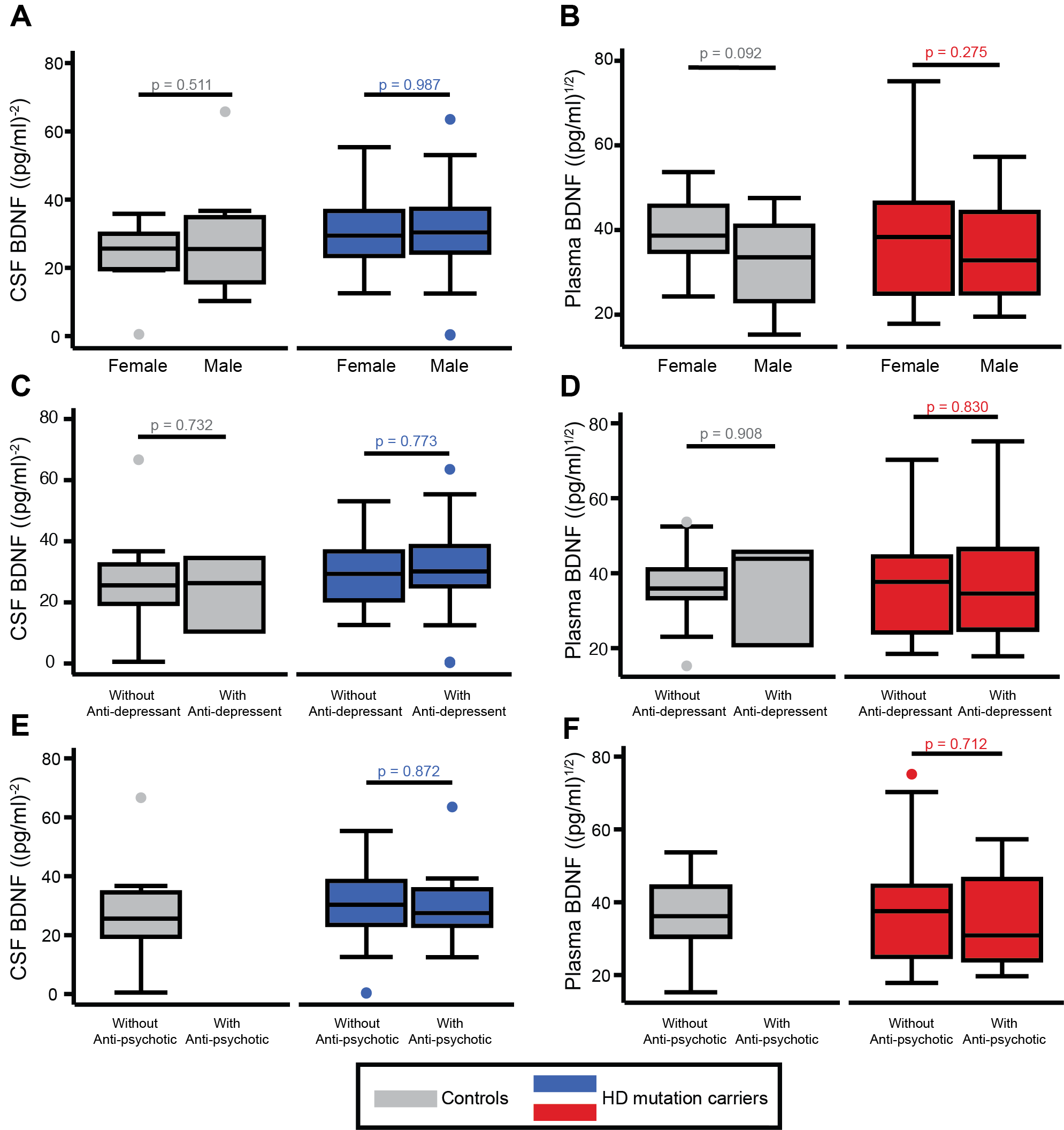

#### **Figure S1. Assessments for potential confounding (categorical variables).**

A boxplot of mean BDNF in CSF and plasma by gender (A,B), anti-depressant users (C,D), and anti-psychotic users (E,F) in controls and HD mutation carriers. Groups were compared using general linear models. For anti-psychotic, only HD mutation carriers were included since healthy controls do not take anti-psychotic medications in our cohort. Results were replicated at follow-up. CSF BDNF values were inverse square transformed and plasma BDNF were square root transformed. CSF, cerebrospinal fluid; BDNF, brain-derived neurotrophic factor.

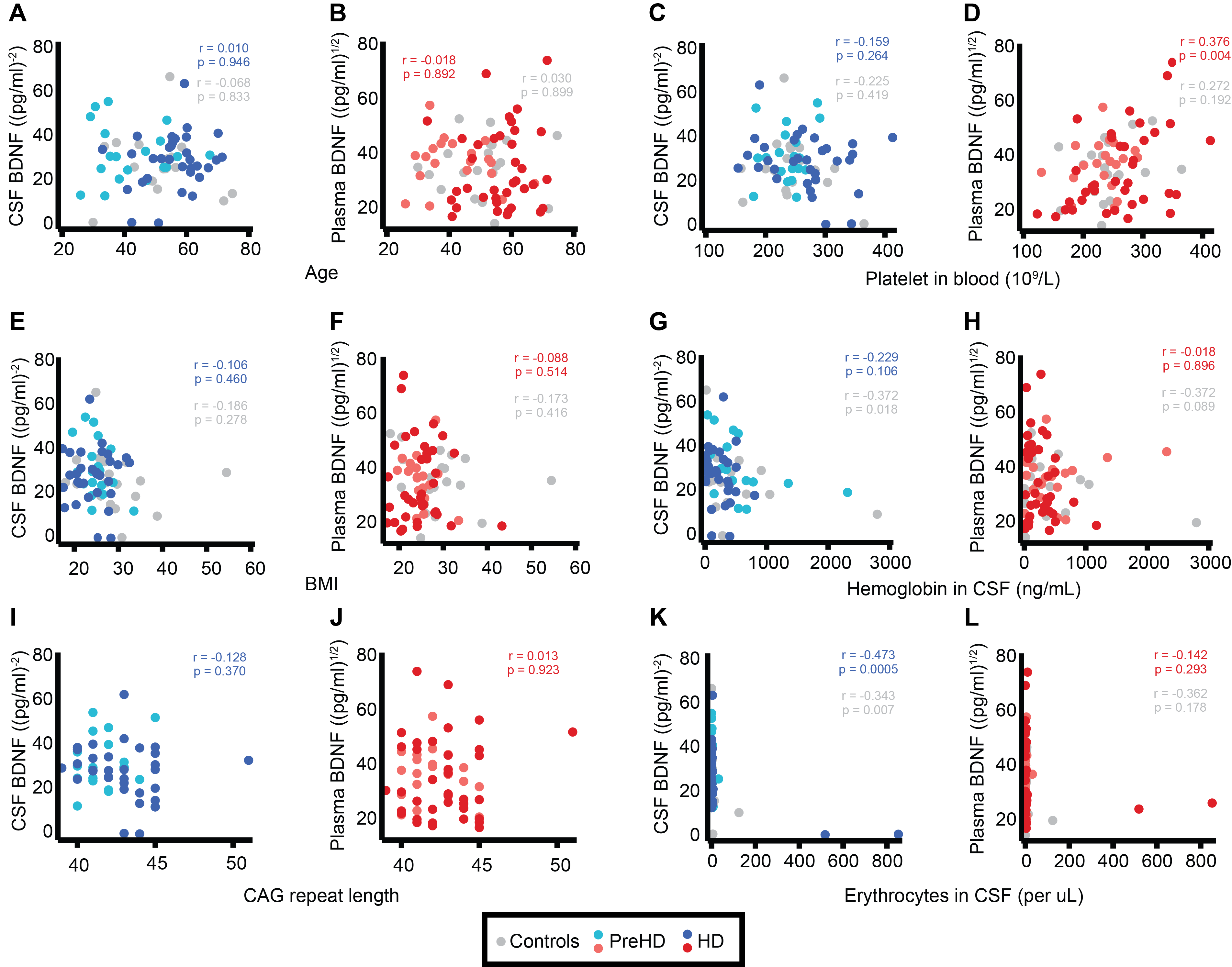

#### **Figure S2. Assessments for potential confounding (continuous variables).**

Relationship between BDNF in CSF (blue; A, C, E, G, I, and K) and plasma (red; B, D, F, H, J, and L) and potential confounding variables. Association were tested using Pearson’s correlations using either healthy controls (n=20, gray data points) and HD mutation carriers (n=57, blue or red data points). For CAG repeat length correlation, only HD mutation carriers were included. Results were replicated at follow-up. CSF BDNF values were inverse square transformed and plasma BDNF were square root transformed. CSF, cerebrospinal fluid; HD, manifest HD mutation carriers; PreHD, premanifest HD mutation carriers; BDNF, brain-derived neurotrophic factor; BMI, body-mass index; CAG, cytosine-adenosine-guanine.

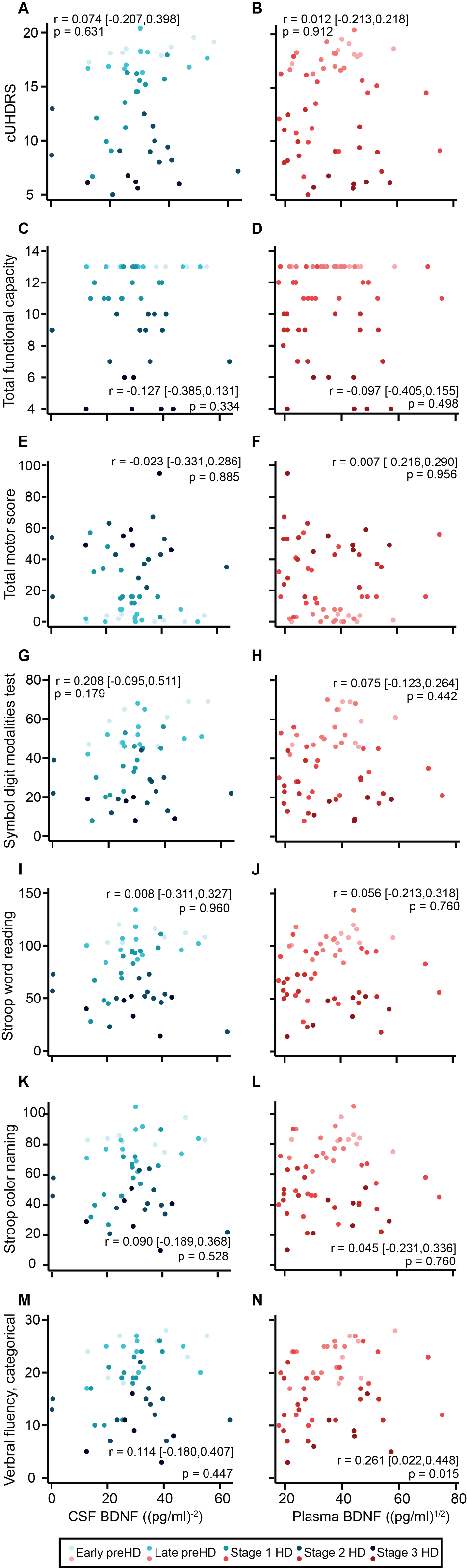

#### **Figure S3. Baseline associations with clinical measures.**

Associations between CSF BDNF (n=51; blue; A, C, E, G, I, and K) and plasma BDNF (n=57; red; B, D, F, H, J, and L) and UHDRS clinical measures (A to D), including TFC (A,B), TMS (C,D), and cognitive measures (E to L), including SDMT (E,F), SWR (G,H), SCN (I,J), and VFC (K, L). Pearson’s partial correlations with adjustments for CSF (gender, BMI, anti-depressant, anti-psychotic medication, age, CAG repeats, and erythrocyte count) or plasma covariates (gender, BMI, anti-depressant, anti-psychotic medication, age, CAG repeats, sample storage duration, and platelet count) and bootstrapped bias corrected accelerated 95% confidence intervals are shown. Results were replicated at follow-up. CSF BDNF values were inverse square transformed and plasma BDNF were square root transformed. CSF, cerebrospinal fluid; HD, manifest HD mutation carriers; PreHD, premanifest HD mutation carriers; BDNF, brain-derived neurotrophic factor; cUHDRS, composite Unified Huntington’s Disease Ratings Scale; TFC, total functional capacity; TMS, total motor score; SDMT, Symbol digit modalities test; SWR, Stroop word reading; SCN, Stroop color naming; VFC, verbal fluency- categorical.

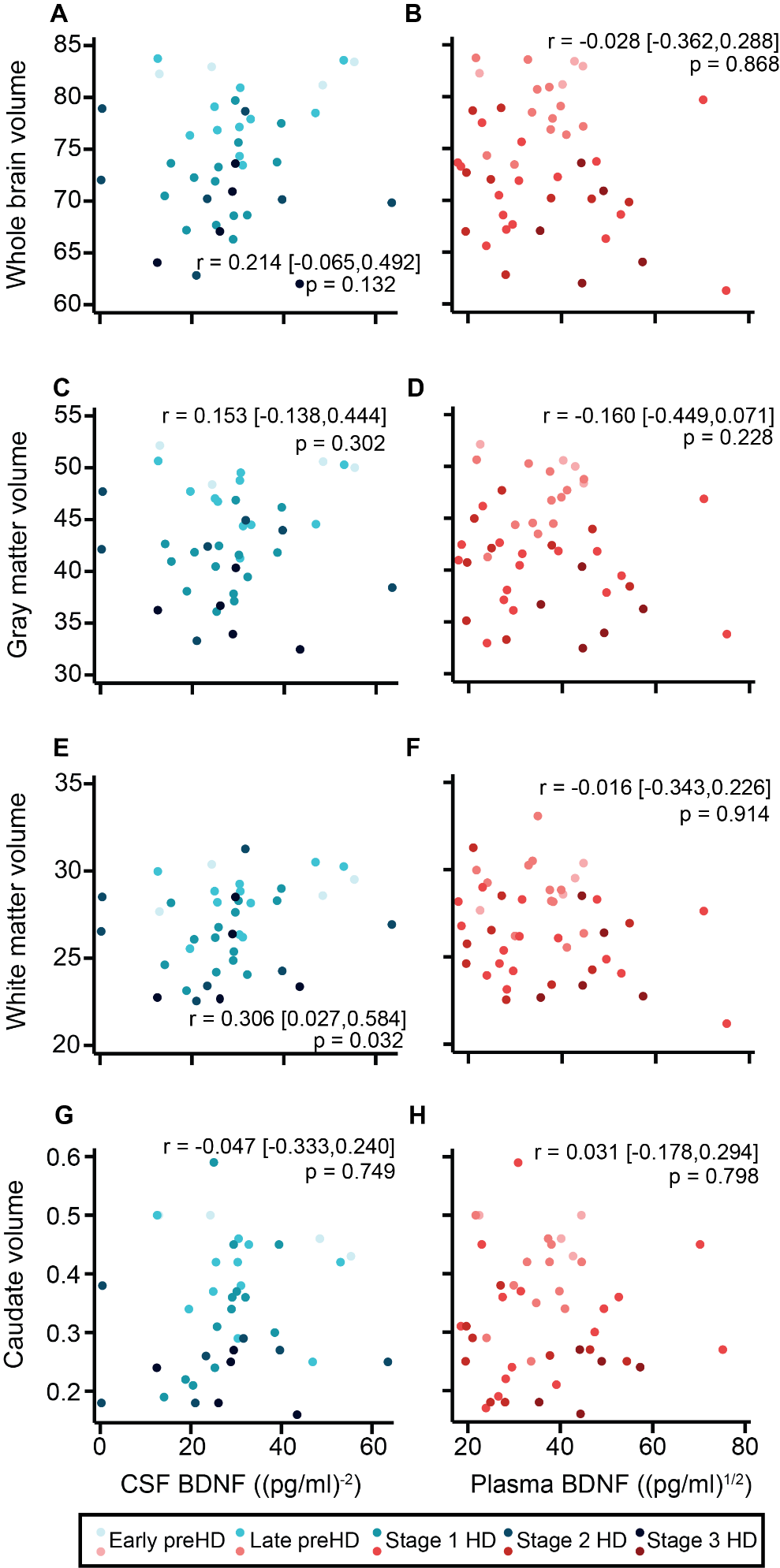

#### **Figure S4. Baseline associations with imaging measures.**

Associations between CSF BDNF (n=41; blue; A, C, E, and G) and plasma BDNF (n=46; red; B, D, F and H) and imaging measures, including MRI volumetric of whole brain (A,B), grey matter (C,D), white matter (E,F), and caudate (G,H). Volumetric measures are percentages of the total intracranial volume (TIV). Pearson’s partial correlations with adjustments for CSF (gender, BMI, anti-depressant, anti-psychotic medication, age, CAG repeats, and erythrocyte count) or plasma covariates (gender, BMI, anti-depressant, anti-psychotic medication, age, CAG repeats, sample storage duration, and platelet count) and bootstrapped bias corrected accelerated 95% confidence intervals are shown. Results were replicated at follow-up. CSF BDNF values were inverse square transformed and plasma BDNF were square root transformed. CSF, cerebrospinal fluid; HD, manifest HD mutation carriers; PreHD, premanifest HD mutation carriers; BDNF, brain-derived neurotrophic factor.

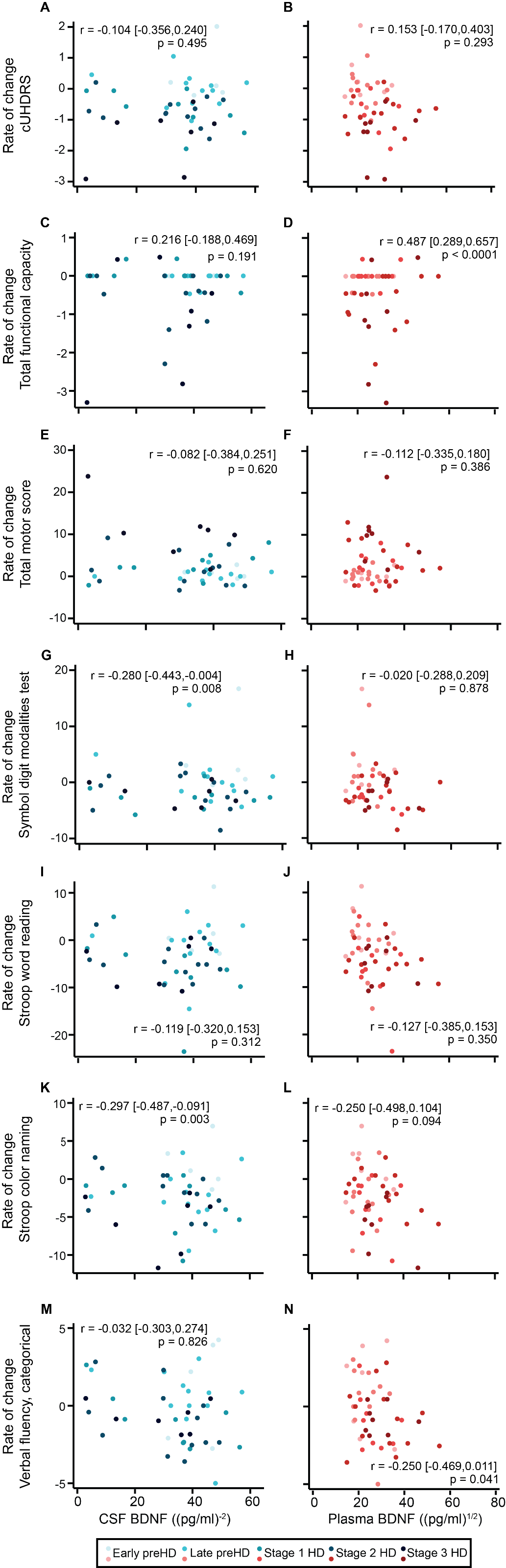

#### **F****igure S5. Associations between baseline BDNF and rate of change in clinical measures.**

Associations between baseline CSF BDNF (n=47; blue; A, C, E, G, I, K, and M) and plasma BDNF (n=52; red; B, D, F, H, J, L, and N) and rate of change UHDRS clinical measures (A-F), including cUHDRS (A,B), TFC (C,D), TMS (E,F), and cognitive measures (G-N), including SDMT (G,H), SWR (I,J), SCN (K,L), and VFC (M,N). Pearson’s partial correlations with adjustments for CSF (gender, BMI, anti-depressant, anti-psychotic medication, age, CAG repeats, and erythrocyte count) or plasma covariates (gender, BMI, anti-depressant, anti-psychotic medication, age, CAG repeats, sample storage duration, and platelet count) and bootstrapped bias corrected accelerated 95% confidence intervals are shown. CSF BDNF values were inverse square transformed and plasma BDNF were square root transformed. CSF, cerebrospinal fluid; HD, manifest HD mutation carriers; PreHD, premanifest HD mutation carriers; BDNF, brain-derived neurotrophic factor; cUHDRS, composite Unified Huntington’s Disease Ratings Scale; TFC, total functional capacity; TMS, total motor score; SDMT, Symbol digit modalities test; SWR, Stroop word reading; SCN, Stroop color naming; VFC, verbal fluency- categorical.

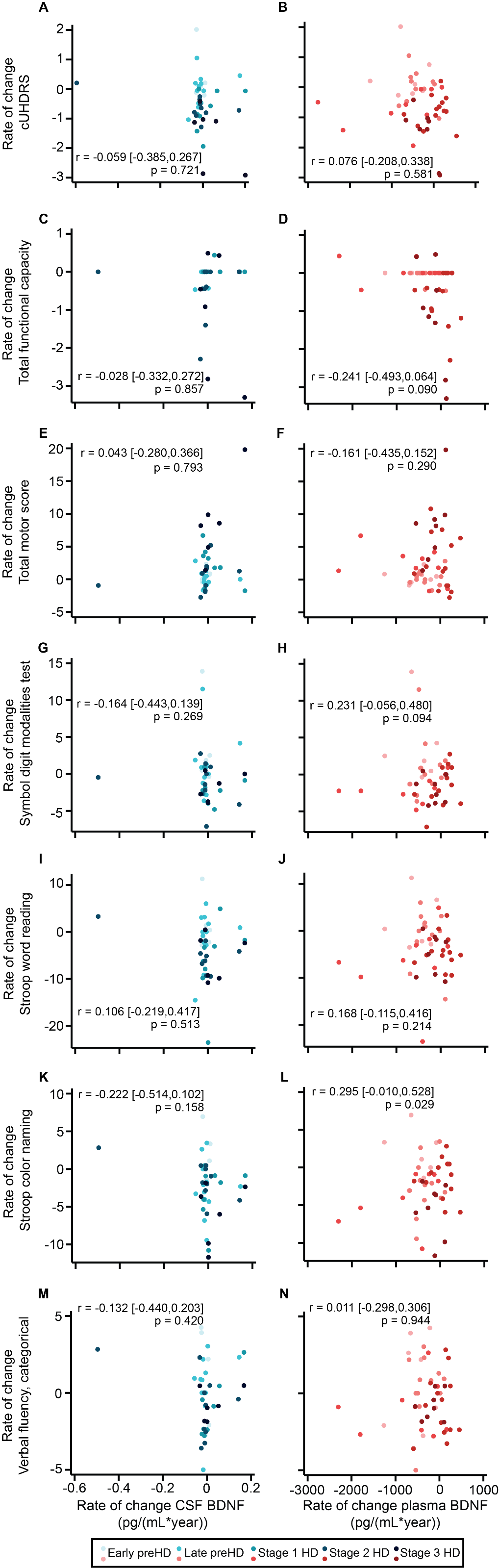

#### **Figure S6. Associations between rate of change in BDNF and clinical measures.**

Association within HD mutation carriers between rate of changes in CSF BDNF (n=43; blue; A, C, E, G, I, K, and M) and plasma BDNF (n=52; red; B, D, F, H, J, L, and N) and rate of change UHDRS clinical measures (A-F), including cUHDRS (A,B), TFC (C,D), TMS (E,F), and cognitive measures (G-N), including SDMT (G,H), SWR (I,J), SCN (K,L), and VFC (M,N). Pearson’s partial correlations with adjustments for CSF (gender, BMI, anti-depressant, anti-psychotic medication, age, CAG repeats, and erythrocyte count) or plasma covariates (gender, BMI, anti-depressant, anti-psychotic medication, age, CAG repeats, sample storage duration, and platelet count) and bootstrapped bias corrected accelerated 95% confidence intervals are shown. CSF BDNF values were inverse square transformed and plasma BDNF were square root transformed. CSF, cerebrospinal fluid; HD, manifest HD mutation carriers; PreHD, premanifest HD mutation carriers; BDNF, brain-derived neurotrophic factor; cUHDRS, composite Unified Huntington’s Disease Ratings Scale; TFC, total functional capacity; TMS, total motor score; SDMT, Symbol digit modalities test; SWR, Stroop word reading; SCN, Stroop color naming; VFC, verbal fluency- categorical
